# Uncontrolled hypertension is associated with an increased risk of end-stage renal disease in patients with type 2 diabetes who underwent postpercutaneous coronary intervention: A nationwide population-based study

**DOI:** 10.1101/2024.04.11.24305701

**Authors:** Hyunglae Kim, Yong-Moon Park, Seung-Hyun Ko, Yun-Jung Cho, Hyung Wook Kim, Yong Kyun Kim, Kyung-Do Han, Jae-Seung Yun, Kyuho Kim, Donggyu Moon

## Abstract

**Background:** Little is known about the use of hypertension to predict end-stage renal disease (ESRD) in patients with type 2 diabetes mellitus (T2DM) and established coronary artery disease who underwent percutaneous coronary intervention (PCI). Here, we evaluated the effect of differential blood pressure (BP) levels on future ESRD in this population.

**Methods:** Using nationwide health check-up data from the Korean National Health Insurance Service between 2015 and 2016, we obtained data for 80,187 patients with T2DM who underwent previous PCI. Patients were classified into four groups according to BP level measured within at least 2 years after PCI: systolic BP <120 (reference), <130, <140, <160, and ≥160 mm Hg; diastolic BP <80 (reference), <90, <100, and ≥100 mm Hg. The primary outcome was incident ESRD, defined as a combination of the relevant disease code and the initiation of renal replacement therapy. Multivariate Cox proportional hazard regression analysis was used to estimate the adjusted hazard ratio (HR) (95% CI) according to BP group.

**Results:** The mean age was 67.7 years, and 80.9% of the participants were treated with antihypertensive medication. ESRD occurred in 1,362 (1.70%) patients during the 4.7-year follow-up. After adjustment for confounding factors, the HR of ESRD patients significantly and sequentially increased in the higher BP groups. Similar findings were noted regarding the relationship between incident ESRD and pulse pressure (PP). According to the subgroup analysis, this relationship was more significant for SBP in those aged <65 years than in those who were aged 65 years or older (*P* for interaction=0.0498). ESRD risk was linearly associated with systolic BP and had a J-shaped association with diastolic BP in patients with baseline values of 70 and 68 mm Hg.

**Conclusions:** In this nationwide population-based study, elevated systolic and diastolic BP and PP were associated with the risk of developing ESRD in a dose‒response manner among T2DM patients who underwent PCI. To prevent ESRD, more strict BP control is needed in T2DM patients who underwent PCI.

## Background

End-stage renal disease (ESRD) is the final phase of chronic kidney disease (CKD) and occurs as a severe chronic consequence of prolonged diabetes, leading to a significant health burden. The incidence of ESRD has been increasing, representing a substantial global public health concern due to the high ESRD-associated rates of morbidity and mortality worldwide. The 2019 Korean Renal Data System report revealed a continuous increase in the incidence and prevalence of ESRD, with diabetes mellitus (DM) being the most common cause of ESRD in Korea (48.4%) ^1^. Although clinical guidelines have been widely disseminated, it is anticipated that the prevalence of ESRD will continue to rise. This is primarily due to the acceleration of societal aging, the exponential rise in chronic metabolic disease, advancements in treatment medications for hypertension and diabetes, and the consequent increase in life expectancy ^1,2^. Hypertension is an important risk factor for cardiovascular disease (CVD) in the general population and among individuals with type 2 diabetes mellitus (T2DM). Regulating blood pressure (BP) within the desired range in individuals with hypertension and/or T2DM decreases the likelihood of cardiovascular incidents and mortality ^3^. Additionally, hypertension is a widely recognized contributing factor in the development and progression of ESRD in individuals with T2DM and in individuals with CKD. Diabetes status is significantly and positively associated with the risk of ESRD. Thus, BP control in individuals with hypertension and T2DM is crucial for preventing ESRD.

In individuals with T2DM, adequate BP control is crucial for reducing the risk of developing CVD, which is the primary cause of mortality and morbidity in these individuals. The latest hypertension guidelines suggest lower BP goals for high-risk patients, including those with CKD or type 2 diabetes. However, the optimal BP target for preventing CKD or its progression in patients with type 2 diabetes and CVD is still unknown. The Korean Society of Hypertension and the Korean Diabetes Association suggest maintaining BP below 140/90 mmHg in people with type 2 diabetes who do not have additional CVD risk factors or CVD. For individuals with T2DM who have one or more risk factors for CVD, existing CVD, CKD stage 3-5, or asymptomatic organ damage, a BP target of less than 130/80 mmHg is advised ^3–5^. However, insufficient data exist on the recommended BP target to reduce progression to ESRD in individuals with coronary artery disease (CAD) and T2DM. Specifically, there are inadequate data on the relationship between BP and ESRD in CAD patients, especially those with T2DM, who have undergone percutaneous coronary intervention (PCI). Using the Korean National Health Insurance Service (KNHIS) database, we examined the relationships of systolic BP (SBP) and diastolic BP (DBP) with the likelihood of ESRD in individuals with T2DM who underwent PCI for CAD to determine the ability of BP to predict ESRD development.

## Methods

### Data sources

In this nationwide cohort study, we used data from the KNHIS database ^6^. The KNHIS database is considered to represent the entire South Korean population, and the details of this database have been previously described ^7,8^. All insured Korean people undergo annual or biennial health examinations that are supported by the KNHIS. The demographic data and all medical expenses for both inpatient and outpatient services, pharmacy dispensing claims, and mortality information are included in the database ^6^. From this dataset, we also collected laboratory test results and questionnaire data corresponding to lifestyle habits.

Initially, 2,612,704 patients with T2DM aged 20 years and older who underwent health checkups between 2015 and 2016 (index year) were identified. Among them, we selected 83,588 patients who had undergone PCI prior to health examination. To avoid confounding associated with preexisting diseases and to minimize the possible effects of reverse causality, we also excluded those who had a history of ESRD before the index year (n=1,499). Finally, after the exclusion of individuals with missing values or missing blood test results in the KNHIS database, the study population ultimately consisted of 80,187 subjects. The study population was followed up until December 31, 2020.

This study was approved by the Institutional Review Board of the Catholic University of Korea St. Vincent’s Hospital (VC23ZISE0366). The need for written informed consent was waived. All methods were performed in accordance with the principles of the Declaration of Helsinki.

### Definition of T2DM, comorbidities, and clinical parameters

The presence of T2DM was defined if any of the following were present: (1) at least one claim per year for a prescription of antidiabetic medication under the International Statistical Classification of Diseases, Tenth Revision (ICD-10) codes E11 to E14 from the claims data or (2) fasting plasma glucose ≥126 mg/dL in the health examination without a prescription for antidiabetic medication ^9,10^. The patients’ PCI status was ascertained using the following procedure codes: M6551-M6554, M6561-M6566, M6572, and M6572 ^11^. Hypertension was defined as a SBP of ≥140 mmHg or DBP of ≥90 mmHg recorded in the health examination or the use of antihypertensive medications combined with the ICD-10 codes (I10-13, I15) ^12^. Dyslipidemia was defined as an ICD-10 code of E78 and a history of lipid-lowering drug use or a total serum cholesterol level ≥240 mg/dL recorded in the health examination database ^13^. CKD was defined as an estimated glomerular filtration rate (eGFR) <60 mL/min·1.73 m^2^, calculated using the CKD Epidemiology Collaboration (EPI) equation ^9^.

A low income was defined as the lowest 20% of socioeconomic status. Body mass index was calculated as the weight (kg) divided by the height (m^2^). Information on current smoking and alcohol consumption was obtained via a questionnaire. Smoking history was categorized as nonsmoker, ex-smoker, or current smoker. Participants were categorized as nondrinker, mild drinker (<30 g/day), or heavy drinker (≥30 g/day) based on alcohol consumption status. Regular exercise was defined as physical activity that was performed at least five times per week ^14^.

Blood samples for the measurement of serum glucose and lipid profiles were drawn after overnight fasting. The quality of the laboratory tests was assessed by the Korean Association for Laboratory Medicine, and the hospitals participating in the NHIS health checkup programs were certified by the NHIS ^14^.

### Measurements and classification of BP levels

After obtaining the anthropometric measurements and following a minimum of 5 min of seated rest, KNHIS data for brachial SBP and DBP were collected using an automatic sphygmomanometer in a sitting position, and the recorded value represents the average of two measurements. Pulse pressure (PP) was calculated by subtracting the DBP value from the SBP value ^7^.

We classified BP into the following categories: SBP <120, 120 to 129, 130 to 139, 140 to 159, and ≥160 mm Hg; DBP <80, 80 to 89, 90 to 99, and ≥100 mm Hg; and PP <40, 40 to 49, 50 to 59, 60 to 69, and ≥70 mm Hg.

We also classified the participants into 5 different BP categories following the 2017 clinical practice guidelines for classifying BP (ref)—(1) normal BP (SBP <120 mm Hg and DBP <80 mm Hg), (2) elevated BP (SBP 120–129 mm Hg and DBP <80 mm Hg), (3) stage 1 hypertension (SBP 130–139 mm Hg or DBP 80–89 mm Hg), (4) low stage 2 hypertension (SBP 140–159 mm Hg or DBP 90–99 mm Hg), and (5) high stage 2 hypertension (SBP ≥160 mm Hg or DBP ≥100 mm Hg)—and evaluated the risk of ESRD in each category using those with normal BP as the reference group.

### Study outcome and follow-up

The primary endpoint was incident ESRD, which was defined using a combination of ICD-10 codes (N18-19, Z49, Z94.0, and Z99.2) and a unique code (V code) that was assigned at the initiation of renal replacement therapy (hemodialysis, V001; peritoneal dialysis, V003) and/or kidney transplantation (V005) during hospitalization ^14^. All medical expenses for dialysis are reimbursed using the Korean Health Insurance Review and Assessment Service database. These patients are also registered as special medical aid beneficiaries. Therefore, we identified every patient with ESRD in the entire South Korean population and analyzed the data for all patients with ESRD who started dialysis. We excluded individuals without previous CKD who had a transplant or dialysis code recorded on the same date as an acute renal failure code. Patients on continuous renal replacement therapy or with acute PD were also excluded ^7,14,15^. The participants were followed up until one of the following occurred: a new diagnosis of ESRD, death, loss of health insurance, or end of the study (December 31, 2020).

### Statistical methods

The baseline characteristics are presented as percentages (standard errors, SEs) for categorical variables and means ± SEs for continuous variables. Nonnormally distributed variables are presented as geometric means (95% CI). Baseline characteristics were compared among the ESRD and other groups using the χ2 test and *t* test. The incidence rates of ESRD are presented per 1,000 person-years. We counted person-years from the date of BP measurement to the date of reported ESRD, loss to follow-up, death, or December 31, 2014, whichever came first. We used multivariable Cox proportional hazards regression models to compute the hazard ratios (HRs) and 95% CIs for the risk of ESRD, using the group with the lowest level of each index as the reference group. We selected potential confounders that were either established risk factors for ESRD or based on prior associations with the risk of ESRD in our study population.

We analyzed associations between BP category and ESRD development using 4 models: Model 1, which was unadjusted; Model 2, which was adjusted for age and sex; Model 3, which was adjusted for Model 2 plus smoking status, alcohol consumption status, physical activity status, income, body mass index, dyslipidemia status, and CKD status; and Model 4, which was adjusted for Model 3 plus fasting glucose levels, antihypertensive medication use, and diabetes duration. The cumulative ESRD incidence was estimated by constructing Kaplan‒Meier curves for the entire follow-up period, and we used the log-rank test to examine differences in ESRD development by BP category.

Restricted cubic spline analysis was used to examine the shape of the relationships between BP indices and the risk of ESRD. We selected the number of knots based on the values of the Akaike information criterion to fit the best approximating model, chose either the first or second knot as a reference, and tested for linearity by the Wald test. Because all BP indices showed linear dose-dependent responses above specific thresholds observed in restricted cubic spline analysis and risk estimates from ordinal BP categories, we calculated HRs and 95% CIs per 10-mm Hg increment of each BP index beyond the level at which the risk linearly increased by restricting the analyses to participants whose BP measurements were above the cutoff values determined from the results. We also performed stratified analysis by age (<65 versus ≥65 years), sex, and the presence of CKD.

All *P* values provided are two-sided, with the level of significance set at <0.05. Statistical analyses were performed using SAS version 9.4 (SAS Institute Inc., Cary, NC, USA).

## Results

### Baseline characteristics

The baseline characteristics of the participants with respect to the development of ESRD are shown in Table 1. The mean age was 67.7 years, and 80.9% of the participants were treated with antihypertensive medications. ESRD occurred in 1,362 (1.70%) patients during the 4.7-year follow-up.

**Table 1.** Baseline characteristics of the study population

|  | ESRD (-) | ESRD (+) | *P*-value |
| --- | --- | --- | --- |
| Number | 78,825 | 1,362 |  |
| Age (years) | 66.8 ± 9.5 | 68.6 ± 8.8 | <0.0001 |
| Age, ≥65 years | 46,512 (59.0) | 924 (67.8) | <0.0001 |
| Sex, Male (%) | 56,542 (71.7) | 934 (68.6) | 0.0104 |
| BMI (kg/m<sup>2</sup>) | 25.1 ± 3.2 | 25.1 ± 3.3 | 0.7134 |
| Waist circumference (cm) | 87.4 ± 8.4 | 88.2 ± 8.9 | 0.0005 |
| Low income (%) | 16,113 (20.4) | 316 (23.2) | 0.0124 |
| Current smoker (%) | 13,618 (17.3) | 173 (12.7) | <0.0001 |
| Alcohol drinker (%) |  |  | <0.0001 |
| None |  |  |  |
| Mild | 19,586 (24.9) | 174 (12.8) |  |
| Heavy | 3,129 (3.97) | 23 (1.69) |  |
| Regular exercise (%) | 17,897 (22.7) | 231 (17.0) | <0.0001 |
| Hypertension medication (%) | 63,571 (80.7) | 1,273 (93.5) | <0.0001 |
| AB | 1,163 (1.48) | 88 (6.46) | <0.0001 |
| ACEi | 10,805 (13.7) | 163 (12.0) | <0.0001 |
| ARB | 43,871 (55.7) | 1,022 (75.0) | <0.0001 |
| Beta-blocker | 29,306 (37.2) | 678 (49.8) | <0.0001 |
| Calcium-channel blocker | 22,738 (28.9) | 808 (59.3) | <0.0001 |
| Diuretics | 8,218 (10.4) | 235 (17.3) | <0.0001 |
| Dyslipidemia (%) | 70,929 (90.0) | 1,202 (88.3) | 0.0352 |
| DM duration (years) |  |  | <0.0001 |
| New onset | 8,300 (10.5) | 44 (3.23) |  |
| < 5 | 17,919 (22.7) | 90 (6.61) |  |
| 5-9 | 17,869 (22.7) | 181 (13.3) |  |
| ≥10 | 34,737 (44.1) | 1,047 (76.9) |  |
| Chronic kidney disease (%) | 16,302 (20.7) | 1,194 (87.7) | <0.0001 |
| Systolic blood pressure (mm Hg) | 127.8 ± 15.6 | 135.7 ± 18.5 | <0.0001 |
| Diastolic blood pressure (mm Hg) | 75.6 ± 10.0 | 76.4 ± 11.4 | 0.0034 |
| Fasting glucose (mg/dL) | 137.5 ± 44.4 | 140.9 ± 61.7 | 0.0057 |
| Estimated GFR (mL/min/1.73 m<sup>2</sup>) | 81.1 ± 59.1 | 38.9 ± 47.7 | <0.0001 |
| Total cholesterol (mg/dL) | 150.0 ± 35.6 | 159.6 ± 44.8 | <0.0001 |
| HDL cholesterol (mg/dL) | 46.7 ± 13.3 | 44.0 ± 13.2 | <0.0001 |
| LDL cholesterol (mg/dL) | 75.6 ± 30.7 | 83.2 ± 35.5 | <0.0001 |
| Triglyceride (mg/dL) | 124.4(124.0-124.9) | 142.2(138.2-146.3) | <0.0001 |

Data are n (%) or mean (SD)
ESRD: end-stage renal disease; BMI: body mass index; ACEi, angiotensin-converting enzyme inhibitors; ARB: Angiotensin II receptor blocker; DM: diabetes mellitus; GFR: glomerular filtration rate; HDL: high density lipoprotein; LDL: low density lipoprotein

Compared to the non-ESRD group, the ESRD group had a greater mean age (68.6 ± 8.8 years vs. 66.8 ± 9.5 years, *P*<0.001), lower income status (23.2% vs. 20.4%, *P=*0.0124), longer duration of diabetes (*P*<0.001), and greater incidence of underlying CKD (87.8% vs. 20.7%, *P*<0.001). Among them, 80.7% of the patients in the non-ESRD group and 93.5% of those in the ESRD group were prescribed hypertension medication, respectively ESRD. The values for mean SBP (127.8 ± 15.6 mm Hg vs. 135.7 ± 18.5 mm Hg, *P*<0.001), mean DBP (75.6 ± 10.0 mm Hg vs. 76.4 ± 11.4 mm Hg, *P=*0.0034), and LDL-C (75.6 ± 30.7 mg/dL vs. 83.2 ± 35.5 mg/dL, *P*<0.001) were greater in the ESRD group than in the non-ESRD group. However, the eGFR (81.1 ± 59.1 mL/min/1.73 m^2^ vs. 38.9 ± 47.7 mL/min/1.73 m^2^, *P*<0.001) was lower in the ESRD group than in the non-ESRD group (Table 1).

### Effects of systolic or diastolic BP on the risk of ESRD in patients with T2DM who underwent PCI

Table 2 shows the multivariable Cox proportional hazard regression analysis to estimate the adjusted hazard ratio (HR) (95% CI) based on the BP groups. The incidence rate of ESRD increased according to BP in both the SBP and DBP categories. SBP <120 mm Hg or DBP <80 mm Hg was considered the reference. After adjustment for covariates, the HRs (95% CIs) of SBP for incident ESRD patients were 1.31 (1.10-1.58), 1.66 (1.40-1.97), 2.71 (2.29-3.20), and 3.92 (3.17-4.84) in the 120-129, 130-139, 140-159, and ≥160 mm Hg categories, respectively. Similar dose‒dependent relationships were also noted between DBP categories and incident ESRD [HRs (95% CI), 1.18 (1.05–1.34), 1.52 (1.27–1.82), and 2.45 (1.89–3.20) in the 80–89, 90–99, and ≥100 mm Hg groups, respectively]. PP remained significantly associated with ESRD risk in a dose-dependent manner after adjusting for multiple variables. When the reference range of PP was defined as <40 mm Hg, the HRs (95% CI) for ESRD development also showed a dose-dependent relationship (Table 2).

**Table 2.** Hazard ratios and 95% confidence intervals for outcomes according to the multivariable Cox proportional regression model

|  | N | IR* | Model 1 | Model 2 | Model 3 | Model 4 |
| --- | --- | --- | --- | --- | --- | --- |
| **SBP** |  |  |  |  |  |  |
| <120 | 21,996 | 2.13 | Reference | Reference | Reference | Reference |
| <130 | 19,592 | 2.70 | 1.26 (1.05-1.52) | 1.25 (1.04-1.50) | 1.35 (1.12-1.62) | 1.31 (1.10-1.58) |
| <140 | 21,280 | 3.38 | 1.58 (1.34-1.88) | 1.55 (1.31-1.83) | 1.73 (1.56-2.05) | 1.66 (1.40-1.97) |
| <160 | 14,372 | 6.09 | 2.86 (2.42-3.37) | 2.72 (2.31-3.21) | 2.88 (2.44-3.40) | 2.71 (2.29-3.20) |
| ≥160 | 2,947 | 10.8 | 5.10 (4.13-6.28) | 4.74 (3.83-5.86) | 4.36 (3.53-5.40) | 3.92 (3.17-4.84) |
| **DBP** |  |  |  |  |  |  |
| <80 | 47,724 | 3.43 | Reference | Reference | Reference | Reference |
| <90 | 24,517 | 3.39 | 0.99 (0.88-1.12) | 1.03 (0.91-1.16) | 1.12 (0.99-1.27) | 1.18 (1.05-1.34) |
| <100 | 6,360 | 4.75 | 1.38 (1.16-1.65) | 1.42 (1.19-1.70) | 1.48 (1.24-1.77) | 1.52 (1.27-1.82) |
| ≥100 | 1,586 | 8.19 | 2.39 (1.84-3.11) | 2.51 (1.93-3.26) | 2.41 (1.85-3.14) | 2.45 (1.89-3.20) |
| **PP** |  |  |  |  |  |  |
| <40 | 6,826 | 1.60 | Reference | Reference | Reference | Reference |
| <50 | 24,404 | 2.35 | 1.47 (1.09-1.98) | 1.44 (1.07-1.74) | 1.47 (1.09-1.99) | 1.45 (1.08-1.96) |
| <60 | 26,805 | 2.87 | 1.80 (1.34-2.41) | 1.73 (1.29-2.32) | 1.82 (1.36-2.44) | 1.70 (1.27-2.29) |
| <70 | 14,565 | 5.00 | 3.13 (2.33-4.20) | 2.94 (2.18-3.95) | 2.95 (2.19-3.97) | 2.60 (1.93-3.50) |
| <80 | 7,587 | 9.78 | 6.14 (4.58-8.25) | 5.67 (4.20-7.65) | 4.99 (3.70-6.72) | 4.26 (3.16-5.74) |

Model 1 was unadjusted.
Model 2 was adjusted for age and sex.
Model 3 was adjusted for Model 2 + smoking, alcohol intake, regular exercise, BMI, income, dyslipidemia, and CKD.
Model 4 was adjusted for Model 3 + antihypertensive medication use, fasting glucose level, and diabetes duration.
\*IR incidence rate, 1,000 person-year
SBP: systolic blood pressure; DBP: diastolic blood pressure; PP: pulse pressure; BMI: body mass index; CKD: chronic kidney disease

In addition, incident ESRD also increased according to hypertension stage. When we defined the reference as SBP <120 mm Hg and DBP <80 mm Hg (normal BP), ESRD risk increased even in the SBP 120-139 mm Hg range. The multivariable-adjusted HRs (95% CIs) for ESRD were 1.32 (1.08-1.62), 1.76 (1.51-2.06), 2.74 (2.30-3.27), and 4.45 (3.37-5.89) in the elevated BP, stage 1, low stage 2, and high stage 2 hypertension groups, respectively, compared to the normal BP (SBP <120 mm Hg and DBP <80 mm Hg) group (Table 3).

**Table 3.**
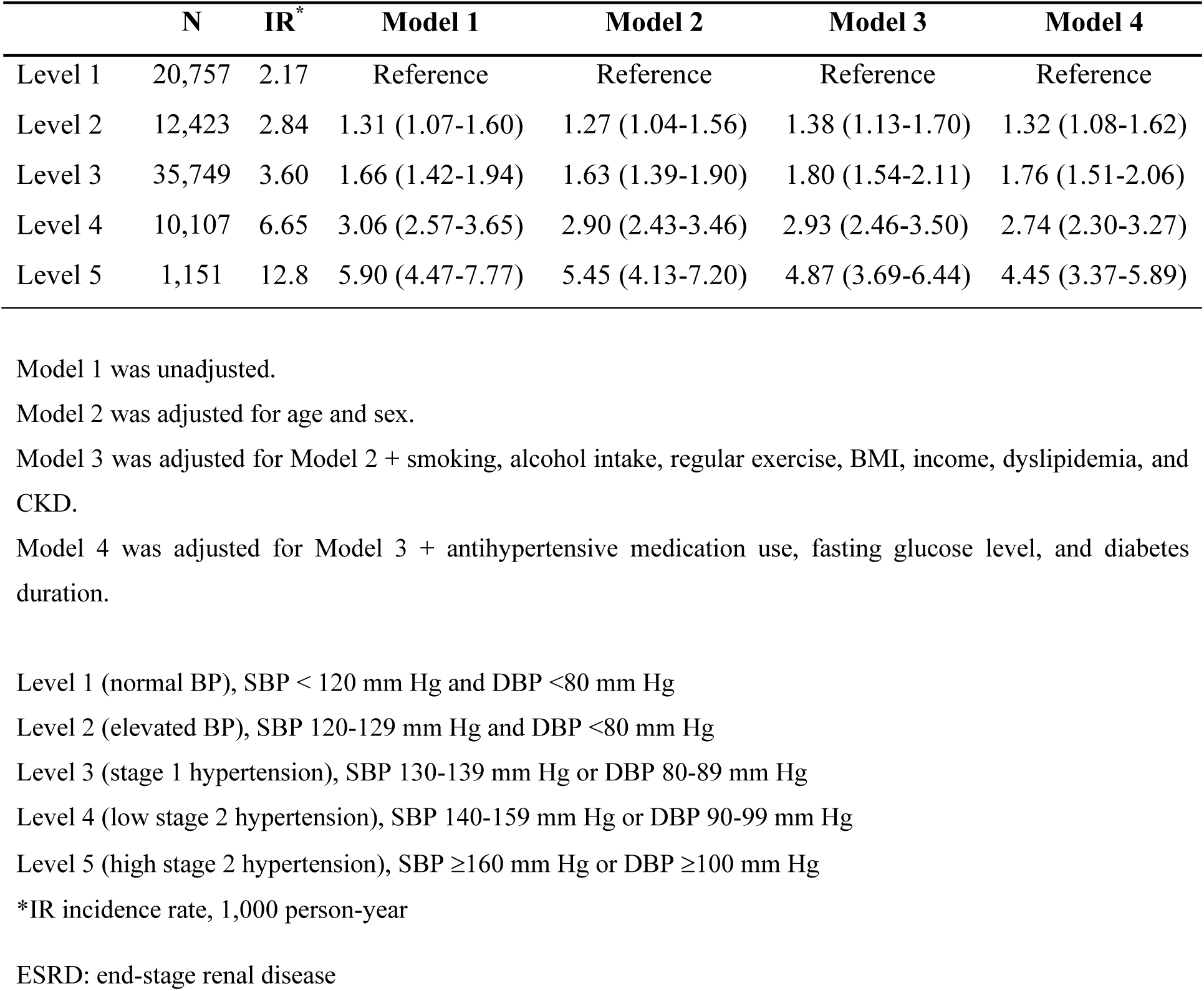
Hazard ratios (95% confidence intervals, CIs) for incident ESRD according to hypertension stage according to the Cox proportional hazards model

We further classified the DBP levels according to SBP categories. In the SBP 140-159 mm Hg group, the lowest DBP group (<80 mm Hg) had the highest HR (95% CI) of 3.20 (2.61-3.92). Moreover, compared to the normal BP group, the highest SBP (≥160 mm Hg) and DBP (≥100 mm Hg) groups had the highest HR (95% CI) (4.86 (3.43-6.88); Table 4).

**Table 4.** Multivariate Cox analysis for incident ESRD according to SBP and DBP in patients who underwent PCI with T2DM

| SBP/DBP<br>(mm Hg) | n | IR | Model 1 | Model 2 | Model 3 | Model 4 |
| --- | --- | --- | --- | --- | --- | --- |
| <120 |  |  |  |  |  |  |
| <80 | 20,757 | 2.17 | 1 (Ref.) | 1 (Ref.) | 1 (Ref.) | 1 (Ref.) |
| 80-89 | 1,178 | 1.41 | 0.65 (0.32, 1.32) | 0.68 (0.34, 1.38) | 0.76 (0.37, 1.53) | 0.83 (0.41, 1.67) |
| 90-99 | 60 | 3.73 | 1.73 (0.24, 12.3) | 1.85 (0.26, 13.2) | 1.68 (0.24, 12.0) | 2.31 (0.32, 16.5) |
| ≥100 | 1 | 0 | - | - | - | - |
| 120-139 |  |  |  |  |  |  |
| <80 | 22,295 | 3.38 | 1.56 (1.31, 1.85) | 1.50 (1.27, 1.78) | 1.61 (1.36, 1.91) | 1.51 (1.27, 1.79) |
| 80-89 | 16,936 | 2.57 | 1.18 (0.97, 1.43) | 1.20 (0.99, 1.45) | 1.36 (1.12, 1.65) | 1.40 (1.15, 1.70) |
| 90-99 | 1,540 | 3.86 | 1.77 (1.20, 2.63) | 1.84 (1.24, 2.73) | 2.10 (1.42, 3.12) | 2.16 (1.45, 3.20) |
| ≥100 | 101 | 2.10 | 0.97 (0.14, 6.88) | 1.02 (0.14, 7.29) | 1.59 (0.22, 11.3) | 1.48 (0.21, 10.6) |
| 140-159 |  |  |  |  |  |  |
| <80 | 4,249 | 8.90 | 4.11 (3.36, 5.02) | 3.79 (3.09, 4.65) | 3.58 (2.92, 4.39) | 3.20 (2.61, 3.92) |
| 80-89 | 5,556 | 5.11 | 2.36 (1.90, 2.93) | 2.27 (1.83, 2.82) | 2.59 (2.08, 3.23) | 2.48 (1.99, 3.09) |
| 90-99 | 3,797 | 4.51 | 2.07 (1.61, 2.68) | 2.06 (1.59, 2.66) | 2.26 (1.74, 2.92) | 2.24 (1.73, 2.90) |
| ≥100 | 770 | 5.82 | 2.68 (1.71, 4.20) | 2.78 (1.77, 4.35) | 2.83 (1.81, 4.43) | 2.81 (1.79, 4.40) |
| ≥160 |  |  |  |  |  |  |
| <80 | 423 | 14.9 | 6.92 (4.66, 10.3) | 6.13 (4.12, 9.12) | 4.88 (3.28, 7.26) | 4.17 (2.80, 6.20) |
| 80-89 | 847 | 12.2 | 5.65 (4.12, 7.75) | 5.20 (3.78, 7.16) | 4.82 (3.51, 6.63) | 4.29 (3.12, 5.91) |
| 90-99 | 963 | 7.25 | 3.34 (2.30, 4.85) | 3.20 (2.21, 4.65) | 3.07 (2.11, 4.45) | 2.74 (1.89, 3.98) |
| ≥100 | 714 | 11.7 | 5.42 (3.84, 7.65) | 5.34 (3.78, 7.55) | 5.05 (3.57, 7.15) | 4.86 (3.43, 6.88) |

Model 1 was unadjusted.
Model 2 was adjusted for age and sex.
Model 3 was adjusted for Model 2 + smoking, alcohol intake, regular exercise, BMI, income, dyslipidemia, and CKD.
Model 4 was adjusted for Model 3 + antihypertensive medication use, fasting glucose level, and diabetes duration.
ESRD: end-stage renal disease; SBP: systolic blood pressure; DBP: diastolic blood pressure; PCI: percutaneous coronary intervention; T2DM: type 2 diabetes mellitus; BMI: body mass index; CKD: chronic kidney disease.

Restricted cubic spline analysis revealed a nonlinear relationship between all BP indices (nonlinearity, P<0.001 for SBP, DBP, and mean arterial pressure (MAP) and P=0.003 for PP) and the risk of ESRD (Figure 1), and the HRs increased in a linear dose-dependent manner beyond a certain level for each index (Figure 1A). The association with the risk of ESRD for SBP appeared to plateau at very low levels, whereas the risk of ESRD seemed to increase with decreasing DBP at very low levels to create an overall J-shaped association (Figure 1B). The risk of ESRD showed a J-shaped association with DBP at 68 mmHg after adjustment for covariates.

**Figure 1.**
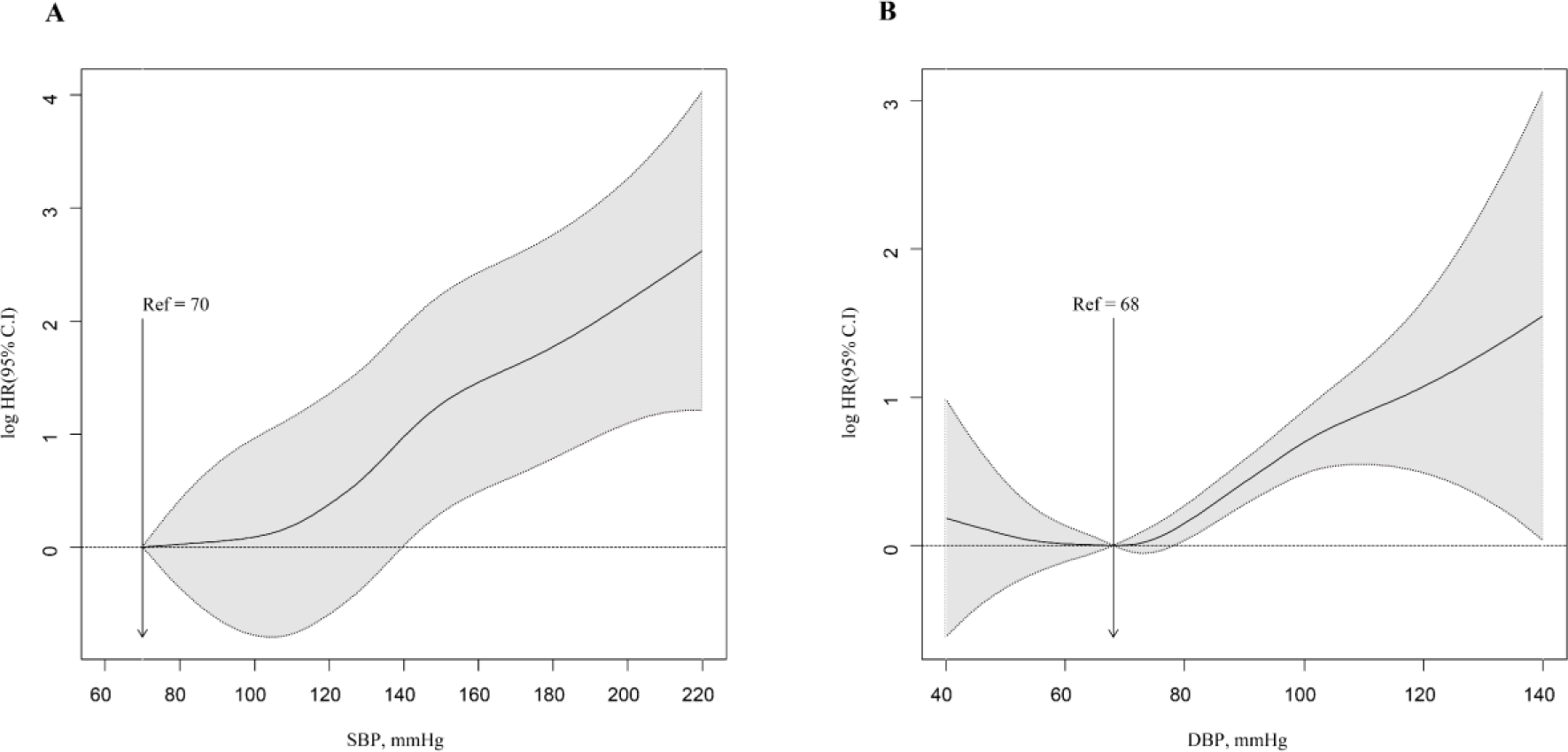
Restricted cubic spine analyses for BP and ESRD patients. The HR for incident ESRD showed a linear relationship with a baseline of 70 mmHg for SBP (A) and a J-shaped curve with a baseline of 68 mmHg for DBP (B). BP: blood pressure; ESRD: end-stage renal disease; SBP: systolic blood pressure; DBP: diastolic blood pressure

### Subgroup Analyses

According to the subgroup analysis based on age, individuals under the age of 65 in the group with a SBP of 160 mmHg or greater exhibited a notably greater HR of 8.368 than did those in the older group, with an HR of 2.741 (P for interaction=0.0498). This finding suggests that the relationship is particularly pronounced in the population under 65 years of age compared to those aged≥65 years (Figure 2). Although not stated here, the risk of ESRD was also linearly correlated with both DBP and PP.

**Figure 2.**
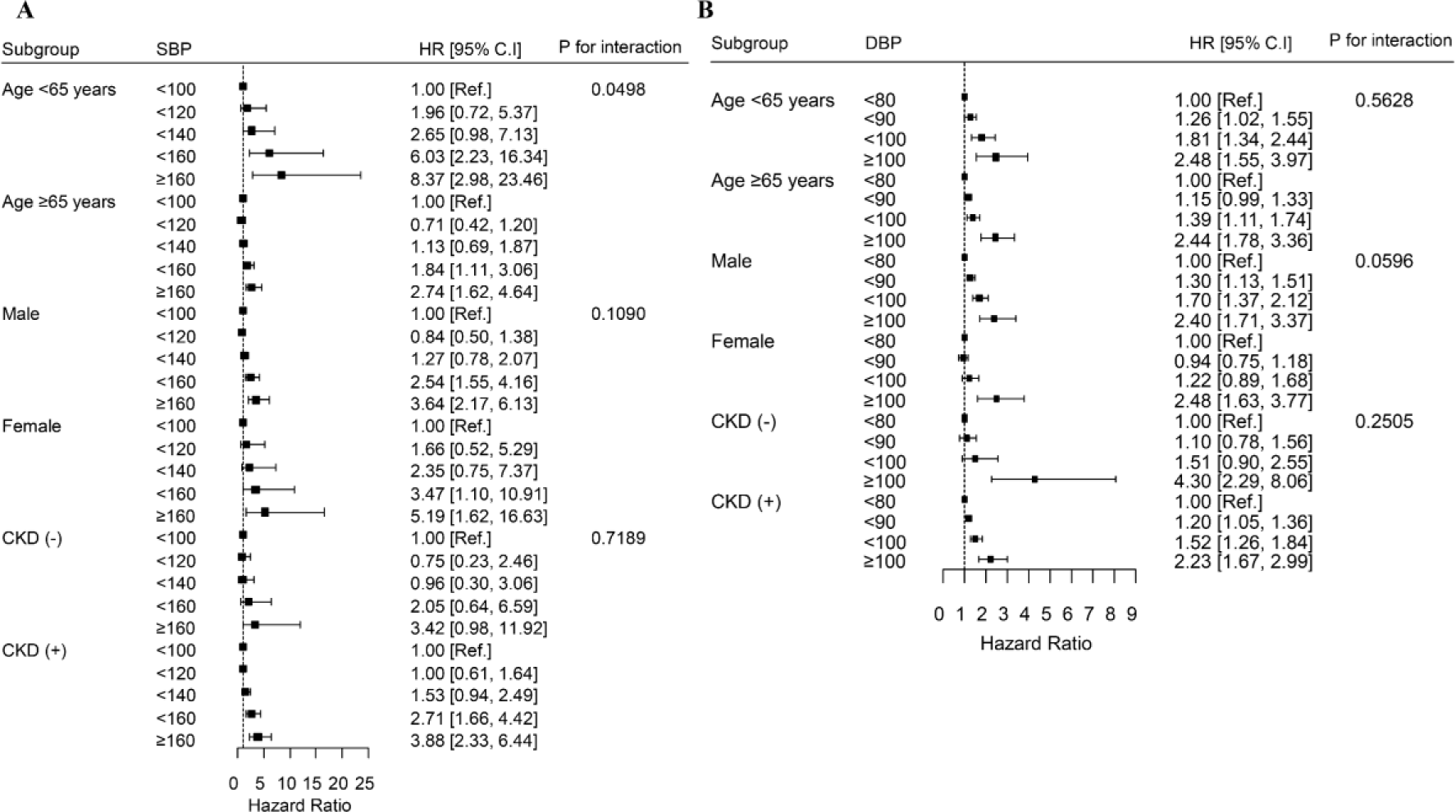
Forest plots for SBP or DBP and ESRD outcomes according to age, sex, and presence of CKD. (A) SBP (B) DBP. SBP: systolic blood pressure; DBP: diastolic blood pressure; ESRD: end-stage renal disease; CKD: chronic kidney disease.

According to the subgroup analysis by sex, compared with male patients (HR=3.644), female patients in the group with a SBP of ≥160 mmHg had a greater risk of ESRD (HR=5.191). According to the subgroup analysis based on history of CKD, the CKD group exhibited a greater HR than the non-CKD group for SBP across all age groups (Figure 2).

## Discussion

We demonstrated direct associations of high SBP, DBP, and PP with the risk of developing ESRD in patients with T2DM who had a history of PCI. Controlling BP within the desired range is crucial for preventing CVD, particularly in individuals with T2DM. Compared with routine treatment, intensive BP lowering was more effective at providing vascular protection ^16^. Previous studies have shown that a decrease of approximately 10 mm Hg in SBP can lead to a 20% reduction in CVD risk, a 28% decrease in heart failure risk, a 27% lower risk of stroke, and a 13% decrease in all-cause mortality ^3^. Furthermore, high-risk patients can have added advantages from more aggressive BP reduction, even if their SBP is less than 140 mmHg ^16^.

It’s as uncertain whether BP should be reduced to the same target level in high-risk groups to prevent ESRD in people with type 2 diabetes. A systematic review and meta-analysis of 19 trials involving 44,989 participants showed that intensive BP-lowering treatment resulted in a 14% (95% CI 4–22) reduction in major cardiovascular events, a 10% (95% CI 3–16) decrease in albuminuria, and a 19% (95% CI 0–34) decrease in retinopathy progression. However, there were no significant impacts on heart failure (15% difference, 95% CI –11 to 34), cardiovascular death (9% difference, 95% CI –11 to 26), or ESRD (10% difference, 95% CI – 6 to 23) (1). Epidemiological studies and randomized controlled trials have shown that BP control is crucial for reducing the decline in eGFR and the risk of future ESRD ^17–20^. However, compared to standard control (less than 140 mm Hg), intensive SBP control (less than 120 mm Hg) in clinical trials did not show any CKD-related benefits in diabetic patients ^21^. Compared to the conventional treatment goal of <140/90 mm Hg, lowering BP to below the current standard of <130/80 mm Hg did not yield additional benefits for overall kidney-related outcomes when in nondiabetic patients with CKD. Moreover, stringent BP management unexpectedly showed a threefold increase in CKD incidence in the group receiving intensive SBP treatment compared to the group receiving conventional treatment ^22^.

Conversely, some research indicates that intensive BP management is crucial for preventing ESRD. Hsu et al. reported that the probability of developing ESRD was greater in patients with diabetes mellitus than in those without DM and that the probability of developing ESRD increased with increasing BP compared to that in individuals with a BP below 120/80 mm Hg^3^. The Kidney Early Evaluation Program (KEEP) study, conducted with a large community-based group of individuals with established CKD, indicated that elevated SBP appeared to be the primary factor contributing to the risk of developing ESRD. However, the increased risk of ESRD was not observable until SBP exceeded 140 mm Hg instead of the recommended target of less than 130 mm Hg, and the increased risk was most significant among individuals with an SBP over 150 mm Hg ^23^. A nationwide population-based cohort study revealed that elevated SBP and DBP were associated with an increased risk of developing ESRD in adult patients with DM. Younger individuals were at particularly higher risk ^15^.

In this investigation, individuals with T2DM who had undergone PCI and had higher BP levels (SBP ≥120 mm Hg, DBP ≥80 mm Hg) exhibited a dose-dependent increase in the risk of ESRD episodes compared to those with normal BP levels. The results observed in the high-risk patient group support the current Korean Diabetes Association (KDA) guidelines recommending a lower BP target (<130/80 mmHg) for hypertensive patients with type 2 diabetes compared to the target for low-risk individuals (<140/90 mmHg). Furthermore, the risk of ESRD was notably elevated even in individuals with BP levels below the hypertensive range (SBP 120-129 mmHg). Specifically, post-PCI DBP exhibited a J-shaped curve starting at a DBP of 68 mm Hg. In individuals with CAD, having a low SBP below 120 mm Hg and a low DBP below 70 mm Hg is linked to a greater likelihood of cardiovascular events, known as the J-curve phenomenon ^24,25^. Since coronary perfusion occurs during the diastolic phase, it has been thought that low diastolic pressure causes coronary perfusion to decrease, which increases the likelihood of cardiac events ^26^. Thus, a target BP of approximately <130/80 mm Hg in patients with CAD appears safe and can thus be recommended; however, a target BP of <120/70 mm Hg is not recommended ^27^.

In this investigation, PP was identified as a significant risk factor for ESRD in this high-risk group. PP is a significant index derived from SBP and DBP to assess the pulsatile element and stiffness of the arteries. PP has been identified as a standalone predictor of cardiovascular mortality and morbidity ^28,29^. The Framingham Heart Study showed that PP was more effective than SBP and DBP in predicting the risk of congestive heart disease ^30^. Furthermore, individuals with elevated PP show an increased likelihood of developing ESRD according to many studies ^30–33^. Within a population-based cohort in Singapore, compared with other BP indicators, such as SBP, DBP, and MAP, PP exhibited the most robust correlation with the risk of ESRD. Arterial stiffness, as indicated by PP, may negatively impact renal function through hemodynamic and metabolic damage ^34–36^. Increased aortic stiffness leads to increased pressure fluctuations and wave reflection, causing elevated pressure and flow fluctuations in the microvascular beds of the kidneys. This can result in microvascular ischemia and injury to renal tissue ^28,37^.

Our study indicates that managing hypertension is crucial for lowering the risk of ESRD. Therefore, enhancing adherence to antihypertensive medication and lifestyle changes are essential beginning in the early stages of CKD ^1^. Less than 45% of individuals with CKD achieve the desired BP targets, and effective management typically involves the use of 3 to 4 drugs ^38,39^. Moreover, the effective rate of BP regulation via antihypertensive medication was approximately 70% among Korean patients with T2DM in 2016, as reported in the Diabetes and Metabolism Journal and the Korean Journal of Internal Medicine ^40^. While there was a rise in hypotension due to more aggressive BP reduction, including severe hypotensive incidents, there was no indication that these negative effects would surpass the advantages of treatment in high-risk patient groups.

Our study has several strengths, including a large sample size, nationwide cohort design, and objective method for detecting ESRD. Nevertheless, our study has several limitations. First, important variables, such as diabetes duration, medication information, combined diabetic complications, and glycemic control status, were not analyzed. Second, we analyzed the results of a one-time BP measurement using different devices during the health examination. However, 1-time BP measurements taken by trained personnel have been shown to correlate with average BP levels, which is an important predictor of adverse outcomes ^41^. Third, there were differences in the time between BP measurement and PCI among the participants. Finally, the population observed in this study consisted of one ethnic group. Despite these limitations, our findings have implications for optimizing the management of hypertension to reduce the risk of ESRD in established CVD patients with T2DM. To mitigate the risk of ESRD, a more strict approach to BP control is needed for patients with T2DM and a history of PCI.

In short, Korean individuals with type 2 diabetes who had previously undergone PCI and had high SBP or DBP had a greater risk of ESRD than did those with normal BP. ESRD is a severe complication for people with longstanding T2DM. Furthermore, individuals with a prolonged history of T2DM frequently experience CVD as a vascular complication. Controlling BP within the target range and providing thorough treatment to prevent ESRD are important considerations in patients with confirmed CVD and T2DM. There was a J-shaped association between DBP and ESRD. However, intensive BP reduction may be advantageous for individuals with documented CAD who have undergone previous PCI. Physicians must proactively manage BP in this high-risk population to prevent ESRD in patients with T2DM. In addition, our findings provide additional evidence that the management of hypertension may require not only reducing SBP and DBP but also maintaining PP for renal protection. Additional research is needed to determine the ideal BP target for individuals with type 2 diabetes at high risk of ESRD.

## Data Availability

Not applicable

## Acknowledgements

Not applicable

## Disclosures

### Ethics approval and consent to participate

This study was approved by the Institutional Review Board of the Catholic University of Korea St. Vincent’s Hospital (IRB no. VC23ZISE0366). The need for written informed consent was waived. All methods were performed in accordance with the principles of the Declaration of Helsinki.

## Authors’ contributions

Conception or design: S.H.K., D.G.M.

Acquisition, analysis, or interpretation of data: K.D.H., Y.M.P. Drafting the work or revising: S.H.K, D.G.M, H.L.K.

Final approval of the manuscript: S.H.K., D.G.M., H.L.K., K.D.H., Y.M.P., J.S.Y., K.H.K., Y.J.C., H.W.K., Y.K.K.

